## Supplemental Table 1 for "Co-design and evaluation of a youth-informed organisational tool to enhance trauma-informed practices in the UK public sector: a study protocol"

SUPPLEMENTARY MATERIALS

This table shows example activities that may be used in the AEBCD series in ways that can be tailored to different ages of young people, and which draw on creative practices and play.

|  | All ages | 10-12 year olds | 13-15 year olds | 16+ year olds |
| --- | --- | --- | --- | --- |
| Showing ‘trigger films’ (standard in EBCD) | Given context for films and purpose in workshop | Asked to spot 1-2 things in short clips (bingo). Given cards to show their view of important / not important / not sure in response to short clips. | Asked to write a word that captures what they felt was important in a clip. Collect and discuss as a group | Open discussion after clips. Encouraged to make brief notes during screening. Can use ‘not sure’ card |
| Sharing data / information | Presented in an engaging way; not using academic terms; giving examples; present as a quiz / myth busting / predicting what the information will say | Using thumbs up / down to indicate if they think it makes sense / is important / worth discussing. | Additionally asked for whether they are surprised by the data (thumbs up / down). Ideas for how we might explain the data / information to someone else. | In small groups, creating Venn diagrams / idea bubbles about what might they notice in the data / information and what might explain it |
| Persona / Character Play: designing a character with a back story to explore and better comprehend the nature of issues / diversity of experience | Given creative materials to design character in small group; asked to answer ‘what has happened to your character’? ‘how did that make them feel?’ and ‘what would they like to happen now’? | Supported to think about how movie characters are made up can still be about real life. Encouraged to focus on physical design of character first and then their story | Given guidance to think first about the big issue / experience they want their character to represent. | Encouraged to think about composite issues that a character could represent |
| Snowball Fight: people write an example on a piece of paper and in groups throw across the room for a set period. Pick up and read out examples near you at end. | Can write or draw the example, word they want to share. | Can ask someone else to read out word on the paper if wished. | Could be invited to think of the best word / example they can | Could be invited to add a brief statement word / example explaining why that is important to them |
| Forum Theatre: inviting or showing a live performance of a scenario, with pauses to ask the ‘audience’ different ways to respond / solve / change the scenario | Show example of how Forum Theatre works and how it can help creativity. Specific boundaries of response s as feasible and realistic in the real world. | Can be offered options if they find it difficult to imagine alternative responses and / or can focus ratings of big / medium / small problems in the scenarios | Supported to try one response option first; given time to consider impact and supported to report what they see. | Could hold opposing response options in mind and discussion in some detail |
| Bullseye Activity: refining the specific aims/ outcomes of a resource or anticipated change | Give examples of the pros and cons of specific vs. general aims, using everyday examples. Practising how we can vote for something as being near or far from our collectively determined bullseye | Invited to design the bullseye page and work with same-age peers to generate ideas for outcomes, using comparisons to everyday examples | Supported to think of ‘wishes’ or ‘if only’ as a way to think about potential change / outcomes. | Supported to consider specific vs general aims via colour coding |
| Implementation mapping: decision trees / if-then steps | Shown examples of mapping resource implementation to understand choices and options, as well as importance of collective clarity | Use of simple ratings (Yes, No, Maybe) on cards for the most important choices. Moving these into piles to help see choices. | Use of ‘must do’, ‘must not do’, ‘not sure’ cards in response to options; adding of red and green flags for point of risk vs opportunity | Supported in groups to sketch starting point and key steps in implementation, considering what would be realistic in their setting. |
